# Unhealthy lifestyle mediates the adverse effect of childhood traumas on acceleration of aging: analysis of 110,596 UK Biobank participants

**DOI:** 10.1101/2022.04.22.22274167

**Authors:** Gan Yang, Xingqi Cao, Xueqin Li, Jingyun Zhang, Chao Ma, Ning Zhang, Qingyun Lu, Eileen M. Crimmins, Thomas M. Gill, Xi Chen, Zuyun Liu

## Abstract

**Background:** Accelerated aging makes adults more vulnerable to chronic diseases and death. This study evaluates the association of childhood traumas with a phenotypic aging measure that captures mortality and morbidity risk, and the role of unhealthy lifestyle in mediating these associations.

**Methods:** We assembled data from 110,596 members of the UK Biobank aged 40-69 years who participated in the baseline survey (2006-2010) and online mental health questionnaire (2016). A phenotypic aging measure—Phenotypic Age Acceleration (PhenoAgeAccel) was calculated, with the higher value indicating the acceleration of aging. Body mass index, smoking status, alcohol consumption, physical activity, and diet were combined to construct an unhealthy lifestyle score (range: 0-5). Childhood traumas including physical neglect, emotional neglect, sexual abuse, physical abuse, and emotional abuse were assessed. General linear regression and formal mediation analysis were performed.

**Results:** Each individual childhood trauma and cumulative childhood traumas were significantly associated with PhenoAgeAccel. For instance, compared with participants who did not experience childhood traumas, those who experienced four (β=0.292, standard error [SE]: 0.091) or five childhood traumas had higher PhenoAgeAccel (β=0.669, SE: 0.169) in fully adjusted models. The formal mediation analysis revealed that unhealthy lifestyle partially mediated the associations of childhood traumas with PhenoAgeAccel (26.1%-42.6%).

**Conclusions:** In a large sample from UKB, childhood traumas were positively associated with acceleration of aging; and more importantly, unhealthy lifestyle partially mediated these associations. These findings reveal a novel pathway from childhood traumas to late-life health through lifestyle and underscore the potential of more psychological strategies beyond lifestyle interventions to promote healthy aging.

## Introduction

Aging is a complex process of multi-system physiological dysregulation, and accelerated aging makes adults more vulnerable to chronic diseases and death [1]. Delaying aging has been the ultimate goal of human beings over a long history. Meanwhile, how to estimate the effects of interventions to delay aging remains a challenge given the lack of comprehensive aging measures. We have developed a novel phenotypic aging measure—Phenotypic Age Acceleration (PhenoAgeAccel)—using nine clinical biomarkers, chosen for their ability to predict mortality and morbidity [2, 3]. This aging measure provides a useful indicator for evaluating effectiveness of geroprotective interventions, identification of risk factors, and elucidation of mechanisms of aging [4].

A number of studies have documented the effect of early life factors such as childhood traumas on late-life health [5-7]. Given that aging serves as the major risk factor of late-life chronic diseases and death, it follows that childhood traumas may accelerate the aging process, and then place persons at higher risk of chronic diseases. A few studies have explored the associations of childhood traumas with telomere length [8] and DNA methylation-based accelerated aging [9][10]. Given that PhenoAgeAccel outperforms aging measures at the molecular level (e.g., telomere length) in predicting adverse health outcomes, it is necessary to evaluate the association of childhood traumas with PhenoAgeAccel.

Of note, how childhood traumas affect accelerated aging remains unknown. Evidence from the recent theoretical and empirical studies suggests that childhood traumas have long-term effects on high-risk behaviors [11], including high-risk HIV behavior, depression, and unhealthy lifestyle, thus, affecting health in late life. For instance, Anda et al. [12] have reported that persons who experienced sexual abuse had nearly three times the odds of smoking compared to those who did not, resulting in high risks of cancer, pulmonary diseases, and other adverse outcomes. Meanwhile, aging is strongly responsive to modifiable lifestyle factors (e.g., smoking, alcohol consumption, physical activities) [13, 14]. Therefore, we hypothesize that unhealthy lifestyles may partially mediate the effect of childhood traumas on accelerating aging, which has not been investigated in previous research.

We conducted this study utilizing data from UK Biobank (UKB), a large population-based cohort study with approximately 500,000 participants aged 40-69 years[15]. This study aimed to examine the associations of childhood traumas with PhenoAgeAccel as well as the role of unhealthy lifestyle as a mediator in the associations.

## Methods

### Study participants

The baseline survey of UKB was conducted in 2006-2010 and 499,309 participants were recruited. In 2016, almost two-thirds of the participants were chosen to conduct an online mental health questionnaire. The 156,749 individuals who participated in both the baseline survey and online mental health survey were eligible for our study [16]. We excluded participants with missing data on childhood traumas (N=3,728), covariates (N=20,702), and PhenoAgeAccel (N=21,723), leaving 110,596 participants for the analysis **(Figure S1)**.

### Assessment of childhood traumas

Childhood traumas, sourced from the 2016 online mental health questionnaire survey, were assessed with five questions representing physical neglect, emotional neglect, sexual abuse, physical abuse, and emotional abuse, using the Childhood Trauma Screener (CTS) [17]:

1. Someone to take to doctor when needed as a child (physical neglect);
2. Felt loved as a child (emotional neglect);
3. Sexually molested as a child (sexual abuse);
4. Physically abused by family as a child (physical abuse);
5. Felt hated by a family member (emotional abuse);

For each question, potential responses included: “never true”, “rarely true”, “sometimes true”, “often”, and “very often true”. Each item was dichotomized as 0 or 1 based on the cut-off points shown in **Table S1** [18]. After inversely scoring physical neglect and emotional neglect, the summary score of five items ranged from 0 to 5, with a higher score denoting more childhood traumas.

### Assessment of lifestyle

As described in previous studies [19], an unhealthy lifestyle score was established by five lifestyle factors, including body mass index (BMI), smoking status, alcohol consumption, drinking status, physical activity, and diet. The data on lifestyle factors were collected by structured questionnaires and 24-hour dietary recall.

According to the recommendations from the World Health Organization, having a BMI lower than 18.5 kg/m^2^ or higher than 24.9 kg/m^2^ was considered as unhealthy. Smoking 100 cigarettes or more in life was classed as ever smoking and thus as unhealthy [19]. According to the guidelines in the UK, the daily consumption of more than one drink for females and more than two drinks for males was considered as unhealthy [19]. For physical activity, engaging in vigorous activity less than 75 minutes or once per week or moderate physical activity less than 150 minutes or 5 days per week was considered as unhealthy [20]. For diet, unhealthy was defined as inadequate intake of more than half of the following components: increased consumption of fruits, vegetables, (shell)fish, dairy products, whole grains, and vegetable oils; reduced or eliminate consumption of refined grains, sugar-sweetened beverages, and unprocessed meats [21].

For each lifestyle factor, an unhealthy level was scored 1 and otherwise, was scored 0. The unhealthy lifestyle score used as a continuous variable was defined as the summary score of 5 lifestyle factors, with a range of 0 to 5. A higher score indicated a higher level of unhealthy lifestyle.

### Phenotypic Age Acceleration

Phenotypic Age (PhenoAge) was developed by regressing the hazard of mortality on forty-two clinical biomarkers and chronological age [2, 3]. Finally, nine clinical biomarkers and chronological age were selected into a parametric proportional hazards model based on the Gompertz distribution and then we converted 10-year mortality risk into units of years. The formula of Phenotypic Age is presented as follows:

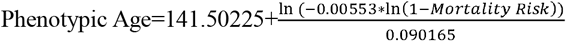

where

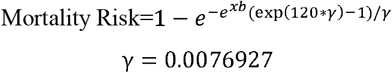

and

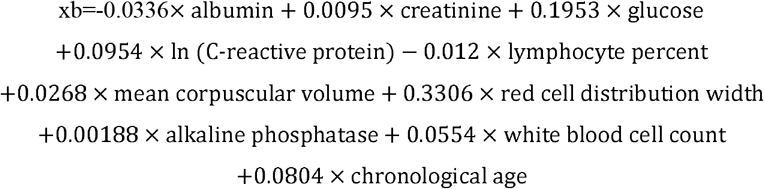

PhenoAgeAccel was calculated as a residual of PhenoAge adjusted for chronological age by linear regression. Participants whose PhenoAgeAccel value was >0 (<0) were defined as phenotypically older (younger). The detailed description of the PhenoAgeAccel has been published elsewhere [2, 3].

### Covariates

Covariates included chronological age, sex, ethnicity, educational level, occupation, Townsend deprivation index (TDI), maternal smoking, and history of cardiovascular disease (CVD) and cancer, which were collected at baseline. Ethnicity was defined as white, mixed, south Asian, black, Chinese, and others [22]. Educational level was classified as high (college or university degree), intermediate (A/AS levels or O levels/GCSEs or equivalent), and low (none of the aforementioned) [23]. Occupation was classified as working, retired, and others (unpaid/voluntary work, full/part time student, looking after home and/or family, unable to work because of sickness or disability, unemployed or did not answer) [22]. TDI utilized census data on employment, housing, and social class based on the postal code of participants [22]. A higher TDI indicated a lower socioeconomic status. Histories of CVD and cancer were classified as yes and no.

### Statistical Analyses

We described the basic characteristics of the participants with mean±standard deviation (SD) for continuous variables and count (percentage) for categorical variables, respectively.

The analytic plan for this study is presented in **Figure 1**. First, general linear regression models were performed to examine the associations of childhood traumas with PhenoAgeAccel. We documented coefficients and corresponding standard errors (SEs) from three models. Model 1 adjusted for sex. Model 2 additionally adjusted for ethnicity, educational level, occupation, TDI, and maternal smoking. Model 3 additionally adjusted for unhealthy lifestyle score.

**Figure 1.**
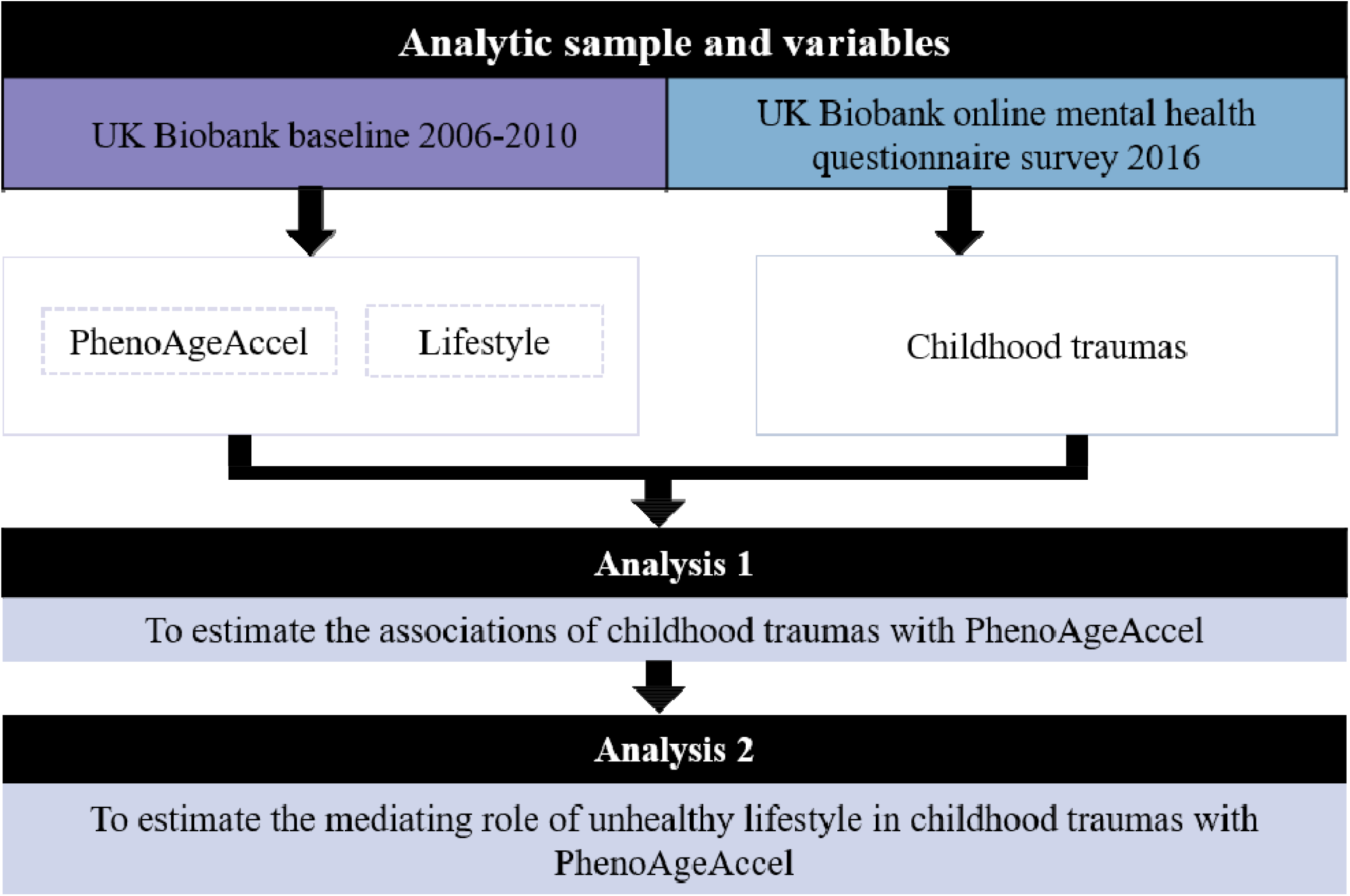
Roadmap for evaluating associations between childhood traumas, unhealthy lifestyle, PhenoAgeAccel. Abbreviations: PhenoAgeAccel: Phenotypic Age Acceleration.

Second, to investigate whether an unhealthy lifestyle mediates the associations of childhood trauma with PhenoAgeAccel, we performed the following analyses in addition to the general linear regression models above. First, general linear regression models were estimated to examine the associations of childhood traumas with unhealthy lifestyle score in two models. Model 1 adjusted for chronological age and sex. Model 2 further adjusted for ethnicity, educational level, occupation, TDI, and maternal smoking. Next, the mediation analysis was performed by R package “mediation” with 1000 simulations. The mediation proportions and corresponding 95% CIs were documented after adjustment for sex. We repeated the above analysis stratified by chronological age (40-59 vs 60-69) and sex.

All analyses were conducted using SAS version 9.4 (SAS Institute, Cary, NC) and R version 4.1.1 (2021-08-10) and P value<0.05 was considered statistically significant.

## Results

### Basic characteristics of participants

The basic characteristics of the participants are presented in **Table 1**. The mean chronological age of the participants was 56.2 (SD=7.7) years. About 56.1% (N=62,017) were females and 97.1% were white. **Figure 2** presents the level and standard error of PhenoAgeAccel in subgroups of childhood traumas. Compared with participants who did not experience childhood traumas, those who experienced physical or emotional neglect or abuse had higher PhenoAgeAccel. The experience sexual abuse was not related to PhenoAgeAccel.

**Table 1.**
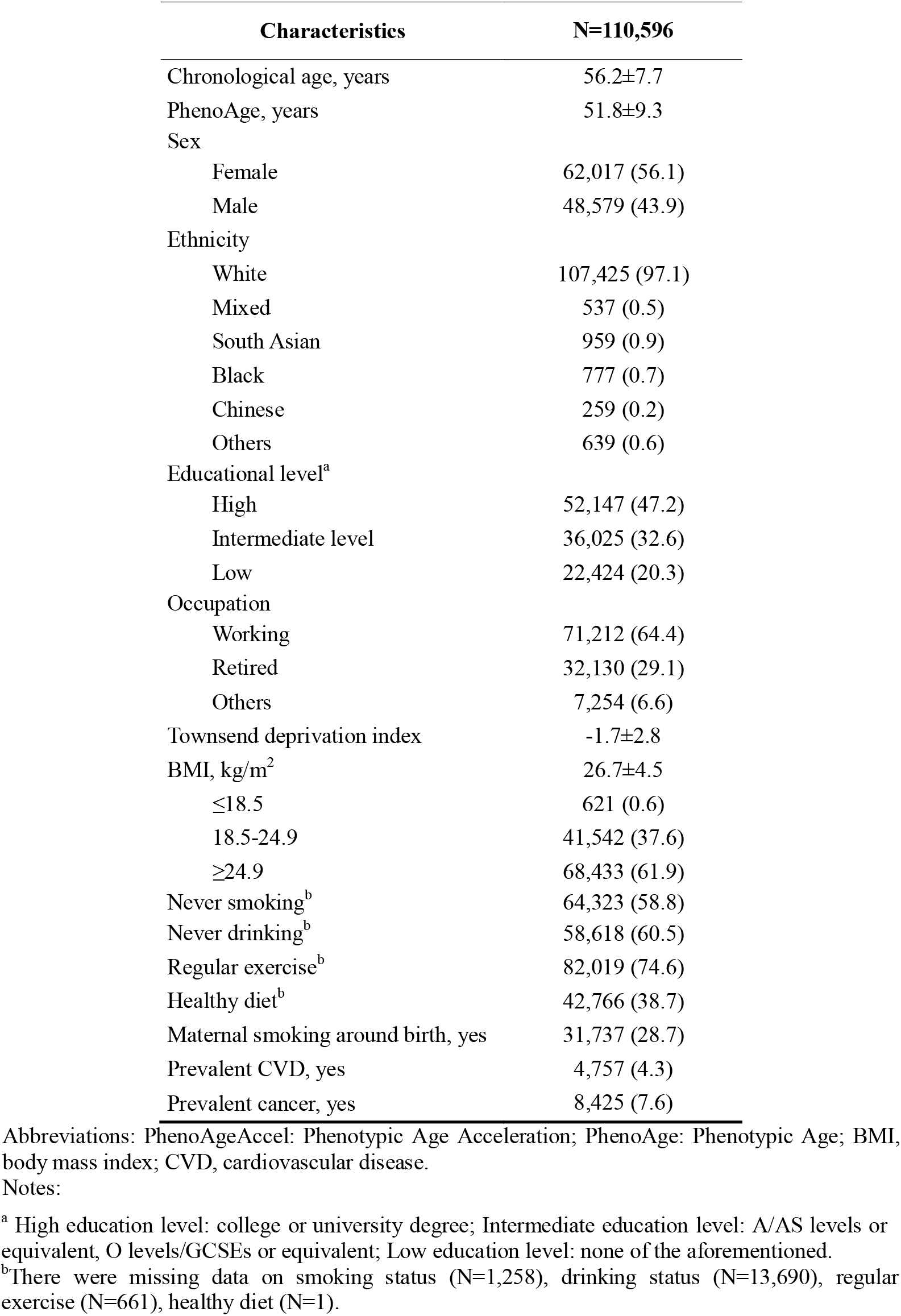
Basic characteristics of the study participants.

| Characteristics | N=110,596 |
| --- | --- |
| Chronological age, years | 56.2±7.7 |
| PhenoAge, years | 51.8±9.3 |
| Sex |  |
| Female | 62,017 (56.1) |
| Male | 48,579 (43.9) |
| Ethnicity |  |
| White | 107,425 (97.1) |
| Mixed | 537 (0.5) |
| South Asian | 959 (0.9) |
| Black | 777 (0.7) |
| Chinese | 259 (0.2) |
| Others | 639 (0.6) |
| Educational level<sup>a</sup> |  |
| High | 52,147 (47.2) |
| Intermediate level | 36,025 (32.6) |
| Low | 22,424 (20.3) |
| Occupation |  |
| Working | 71,212 (64.4) |
| Retired | 32,130 (29.1) |
| Others | 7,254 (6.6) |
| Townsend deprivation index | -1.7±2.8 |
| BMI, kg/m<sup>2</sup> | 26.7±4.5 |
| ≤18.5 | 621 (0.6) |
| 18.5-24.9 | 41,542 (37.6) |
| ≥24.9 | 68,433 (61.9) |
| Never smoking<sup>b</sup> | 64,323 (58.8) |
| Never drinking<sup>b</sup> | 58,618 (60.5) |
| Regular exercise<sup>b</sup> | 82,019 (74.6) |
| Healthy diet<sup>b</sup> | 42,766 (38.7) |
| Maternal smoking around birth, yes | 31,737 (28.7) |
| Prevalent CVD, yes | 4,757 (4.3) |
| Prevalent cancer, yes | 8,425 (7.6) |

Abbreviations: PhenoAgeAccel: Phenotypic Age Acceleration; PhenoAge: Phenotypic Age; BMI, body mass index; CVD, cardiovascular disease.
Notes:
<sup>a</sup> High education level: college or university degree; Intermediate education level: A/AS levels or equivalent, O levels/GCSEs or equivalent; Low education level: none of the aforementioned.
<sup>b</sup>There were missing data on smoking status (N=1,258), drinking status (N=13,690), regular exercise (N=661), healthy diet (N=1).

**Figure 2.**
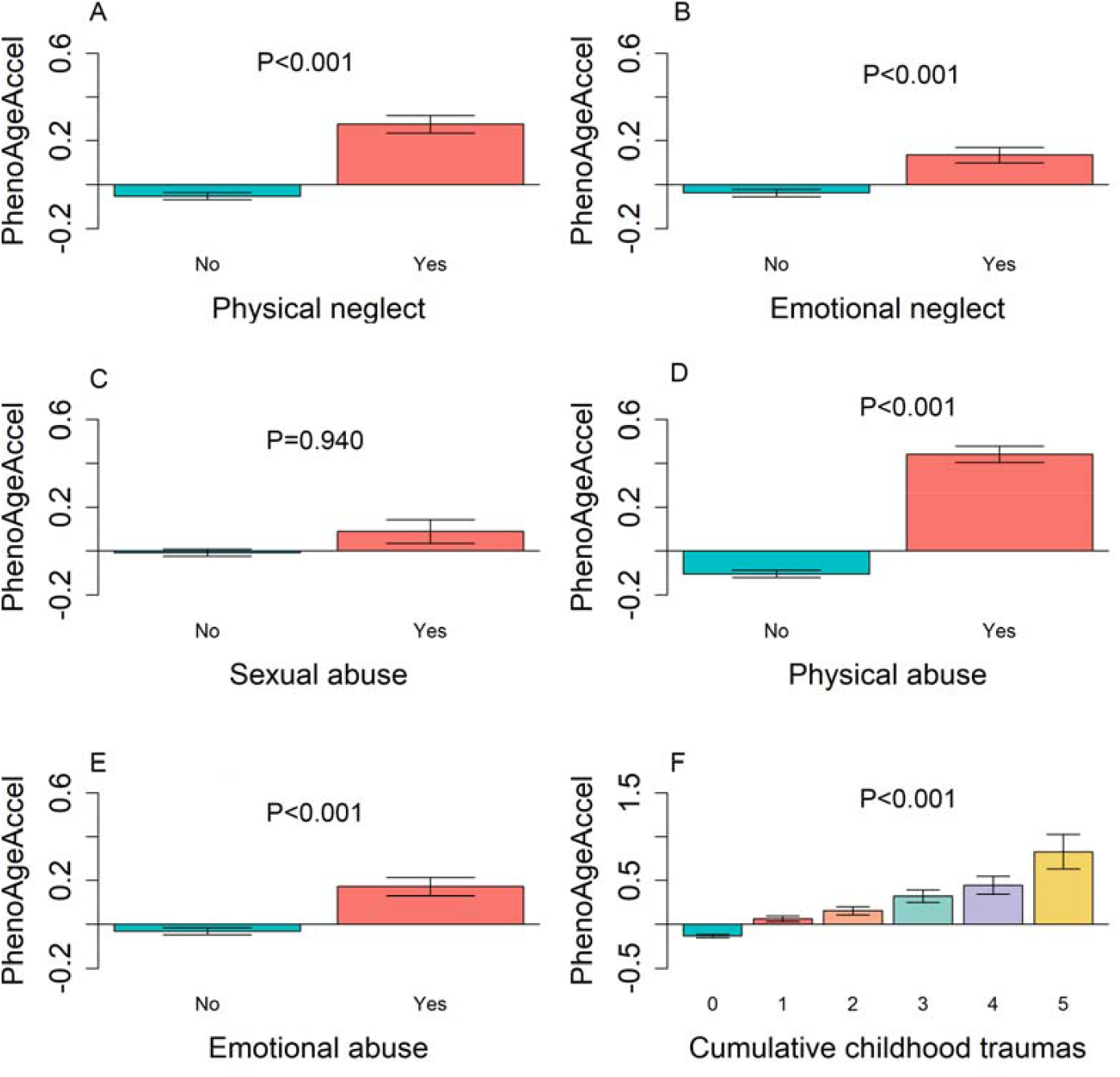
PhenoAgeAccel in subgroups of childhood traumas. **A** PhenoAgeAccel in subgroups of physical neglect; **B** PhenoAgeAccel in subgroups of emotional neglect; **C** PhenoAgeAccel in subgroups of sexual abuse; **D** PhenoAgeAccel in subgroups of physical abuse; **E** PhenoAgeAccel in subgroups of emotional abuse; **F** PhenoAgeAccel in subgroups of cumulative childhood traumas. Abbreviations: PhenoAgeAccel: Phenotypic Age Acceleration.

### Associations of childhood traumas with PhenoAgeAccel

**Table 2** shows significant associations of childhood traumas with PhenoAgeAccel. For instance, compared with participants who did not experience any childhood trauma, those who experienced physical neglect had a significant increase in PhenoAgeAccel (β=0.401, SE: 0.042) after adjusting for sex (Model 1). After further adjustment for more covariates including ethnicity, educational level, occupation, TDI, maternal smoking around birth, and history of CVD and cancer, these associations were maintained (Model 2). The strength of these associations was largely reduced but maintained statistically significant after additionally adjusting for the unhealthy lifestyle score (Model 3). Also, for cumulative childhood traumas, we observed significant dose-response associations (all P for trend<0.05). In the fully adjusted model, participants who experienced four (β=0.292, SE: 0.091) or five childhood traumas had higher PhenoAgeAccel (β=0.669, SE: 0.169), compared with those who did not experience traumas.

**Table 2.** Associations of childhood traumas with PhenoAgeAccel and mediation proportion of childhood traumas in PhenoAgeAccel attributed to unhealthy lifestyle.

| Childhood traumas | Model 1 |  | Model 2 |  | Model 3 |  | Mediation proportion (%) (95% CI)<sup>a</sup> | P value |
| --- | --- | --- | --- | --- | --- | --- | --- | --- |
|  | β (SE) | P value | β (SE) | P value | β (SE) | P value |  |  |
| Physical neglect | 0.401 (0.042) | <0.001 | 0.222 (0.042) | <0.001 | 0.205 (0.041) | <0.001 | 15.1 (11.4, 19.0) | <0.001 |
| Emotional neglect | 0.203 (0.037) | <0.001 | 0.088 (0.037) | 0.017 | 0.009 (0.036) | 0.797 | 55.1 (40.8, 87.0) | <0.001 |
| Sexual abuse | 0.352 (0.054) | <0.001 | 0.260 (0.054) | <0.001 | 0.149 (0.053) | 0.005 | 40.4 (30.6, 55.0) | <0.001 |
| Physical abuse | 0.430 (0.039) | <0.001 | 0.315 (0.039) | <0.001 | 0.208 (0.038) | <0.001 | 32.4 (27.6, 41.0) | <0.001 |
| Emotional abuse | 0.347 (0.042) | <0.001 | 0.229 (0.042) | <0.001 | 0.129 (0.042) | 0.002 | 38.3 (30.5, 51.0) | <0.001 |
| Cumulative childhood traumas (0-5) | 0.184 (0.014) | <0.001 | 0.118 (0.014) | <0.001 | 0.074 (0.014) | <0.001 | — | — |
| 0 | Ref. | — | Ref. | — | Ref. | — | — | — |
| 1 | 0.164 (0.036) | <0.001 | 0.099 (0.036) | 0.006 | 0.061 (0.036) | 0.089 | 35.1 (23.8, 59.0) | <0.001 |
| 2 | 0.291 (0.049) | <0.001 | 0.167 (0.049) | 0.001 | 0.079 (0.049) | 0.107 | 42.6 (32.2, 63.0) | <0.001 |
| 3 | 0.544 (0.067) | <0.001 | 0.357 (0.067) | <0.001 | 0.224 (0.066) | 0.001 | 33.7 (26.4, 45.0) | <0.001 |
| 4 | 0.727 (0.092) | <0.001 | 0.465 (0.092) | <0.001 | 0.292 (0.091) | 0.001 | 33.1 (25.6, 44.0) | <0.001 |
| 5 | 1.319 (0.169) | <0.001 | 0.914 (0.169) | <0.001 | 0.669 (0.167) | <0.001 | 26.1 (19.6, 35.0) | <0.001 |

Abbreviations: SE, standard error; PhenoAgeAccel: Phenotypic Age Acceleration.
Notes:
Model 1: adjusted for sex.
Model 2: further adjusted for ethnicity, educational level, occupation, Townsend deprivation index, maternal smoking around birth, and history of cardiovascular disease and cancer based on Model 1.
Model 3: further adjusted for unhealthy lifestyle score based on Model 2.
<sup>a</sup>The model included sex, and unhealthy lifestyle score.

### Mediation analyses of unhealthy lifestyle in the association of childhood traumas with PhenoAgeAccel

First, the association between childhood traumas and unhealthy lifestyle is shown in **Table 3**. In the fully adjusted model (i.e., Model 2), childhood traumas and cumulative traumas were positively associated with unhealthy lifestyle score. For instance, compared with participants who did not experience any childhood traumas, those who experienced four (β=0.271, SE: 0.195) or five childhood traumas had an increased unhealthy lifestyle score (β=0.412, SE: 0.035).

**Table 3.** Associations of childhood traumas with unhealthy lifestyle score.

| Childhood traumas | Model 1 |  | Model 2 |  |
| --- | --- | --- | --- | --- |
| | $\beta$ (SE) | P value | $\beta$ (SE) | P value |
| Physical neglect | 0.082 (0.009) | <0.001 | 0.023 (0.009) | 0.008 |
| Emotional neglect | 0.160 (0.008) | <0.001 | 0.121 (0.008) | <0.001 |
| Sexual abuse | 0.207 (0.011) | <0.001 | 0.180 (0.011) | <0.001 |
| Physical abuse | 0.204 (0.008) | <0.001 | 0.169 (0.008) | <0.001 |
| Emotional abuse | 0.191 (0.009) | <0.001 | 0.159 (0.009) | <0.001 |
| Cumulative childhood traumas (0-5) | 0.090 (0.003) | <0.001 | 0.070 (0.003) | <0.001 |
| 0 | Ref. | — | Ref. | — |
| 1 | 0.087 (0.008) | <0.001 | 0.065 (0.008) | <0.001 |
| 2 | 0.180 (0.011) | <0.001 | 0.140 (0.010) | <0.001 |
| 3 | 0.256 (0.014) | <0.001 | 0.200 (0.014) | <0.001 |
| 4 | 0.350 (0.020) | <0.001 | 0.271 (0.195) | <0.001 |
| 5 | 0.520 (0.036) | <0.001 | 0.412 (0.035) | <0.001 |

Abbreviations: SE, standard error.
Notes:
Model 1: adjusted for chronological age, and sex
Model 2: further adjusted for ethnicity, educational level, occupation, Townsend deprivation index, and maternal smoking around birth based on Model 1.

Second, the association of unhealthy lifestyle with PhenoAgeAccel is presented in **Table S2**. With a 1-point increase in unhealthy lifestyle score, PhenoAgeAccel increased by 0.69 (SE: 0.039) (Model 1). After adjusting for other covariates, the results remained unchanged in model 2.

Finally, as shown in **Table 2**, an unhealthy lifestyle partially mediated the associations of childhood traumas with PhenoAgeAccel. Relative to participants experiencing none of childhood traumas, unhealthy lifestyle partially mediated 26.1%-42.6% of PhenoAgeAccel for those who experienced some cumulative childhood traumas.

### Subgroup analyses

First, **Table S3** shows the analyses stratified by chronological age. The associations were robust and there were no interactions. Second, the stratification analyses showed significant interactions of some individual and cumulative childhood traumas with sex on PhenoAgeAccel (**Table S4**). For example, the associations of cumulative childhood traumas with PhenoAgeAccel were more pronounced in males (P for interaction<0.001).

## Discussion

In this large sample of over 110,000 adults aged 40-69 years in the UKB, we found that childhood traumas were significantly associated with acceleration of Phenotypic aging. More importantly, we demonstrated that unhealthy lifestyle partially mediated the associations. The findings highlight the importance of reducing traumatic experiences in early life. Furthermore, the findings reveal a novel pathway linking childhood traumas to aging and suggest the potential of lifestyle interventions as well as other strategies to slow aging among adults who have already experienced childhood traumas.

Few studies have explored the association between childhood traumas and PhenoAgeAccel. Our previous study conducted in the US population found that childhood traumas contributed to partial variance in PhenoAgeAccel, which was consistent with the findings of this study [24]. The robust findings of the associations between childhood traumas and Phenotypic aging have important public implications for interventions on adverse childhood experiences in early life to improve health and diminish health inequality in late life. Not just the family and school, but also the whole community and society should closely track the dynamics of physical and emotional adversity in young teenagers.

For the first time, our study demonstrated that unhealthy lifestyle partially mediated the associations of childhood traumas with PhenoAgeAccel, providing clues for mechanisms linking childhood traumas to aging. Prior studies suggested that childhood traumas get under the skin in at least two ways [25-27]. Childhood traumas put individuals under persistent chronic stress and lead to the inequity in individual susceptibility to stress [25]. The cumulative responses to stress produce wear and tear on the system, that is, allostatic load. Allostatic load is a status of cumulative physiological dysregulation accelerating aging and predicting relatively higher risk of mortality and morbidity in older adults [28]. In addition, individuals who suffered childhood traumas might be more likely to adopt unhealthy lifestyles which are socially patterned (e.g., poor diet, smoking, drinking) as a way to avoid or reduce stress which may potentially lead to phenotypic aging in the long term [29]. In fact, childhood traumas have been associatedwith a high risk of alcoholism, smoking, physical inactivity, and severe obesity [30]. Several studies have demonstrated that adherence to a healthy lifestyle may slow phenotypic aging [31-35]. This confirms our hypothesis, childhood traumas → unhealthy lifestyle → accelerated phenotypic aging. Our findings extend previous studies on individual lifestyle factors such as smoking or other substance use behaviors, which have been shown to mediate the association between childhood poverty-related stress and allostatic load [28]. Furthermore, the findings of mediation analysis highlight the importance of lifestyle interventions to mitigate accelerated aging process. More importantly, the partial mediation suggests that among adults who have already experienced childhood traumas, more programs such as stress-resistant strategies are needed beyond lifestyle intervention.

The consistent results of childhood traumas with PhenoAgeAccel in population subgroups strengthen our findings. Of note, we found a significant interaction between cumulative childhood traumas on PhenoAgeAccel stratified by sex. Males seemed to be more susceptible to cumulative childhood traumas. This is complex given the sex differences in hormones, psychology, developmental speed [36] and further studies focused on sex differences are needed in the future.

The present study has some strengths including the large sample size of middle-age and older adults and a series of stratification analyses that were performed to confirm the validity of the findings. This study also has limitations. First, older adults may not accurately recall childhood experiences, resulting in age-dependent memory bias. However, research with prospective reports of childhood traumas, which may be more accurate have demonstrated similar results [37]. Second, our study did not assess the severity and the duration of childhood traumas. In moving forward, further studies should consider multiple aspects of traumas, including the age when traumas first occurred, to reinforce the findings of our study. Third, potential survivor bias may exist. Fourth, most participants in UKB are White and had high socioeconomic status, which may introduce selection bias and potential confounders.

## Conclusions

In summary, among adults aged 40-69 years old in UKB, childhood traumas were associated with acceleration of aging. Furthermore, unhealthy lifestyle partially mediated the associations. The findings call for more attention to childhood traumas in young teenagers. More importantly, the findings reveal a novel pathway linking childhood traumas to late-life health through aging and underscore the potential of lifestyle intervention as well as other strategies to promote healthy aging among adults who have already experienced childhood traumas.

## Data Availability

The UK Biobank data are available online at https://biobank.ndph.ox.ac.uk/showcase/. This study was based on the Application 61856 in UK Biobank.

## Acknowledgments

This research has been conducted using the UK Biobank resource under application number 61856. We wish to acknowledge the UK Biobank participants who provided the sample that made the data available.

